# Naloxone administration and toxicity signs and symptoms: An observational study using Canadian death investigation data

**DOI:** 10.64898/2026.09.04.26362234

**Authors:** Max Ferguson, Claire Healey, Aaron Orkin, Matthew Bowes, Graham Jones, Amanda VanSteelandt

**Author notes:** **Corresponding Author:** (AV).

## Abstract

A toxic drug supply is responsible for a high number of deaths in Canada. Naloxone is a medication that can temporarily reverse opioid toxicity but cannot act on other drug classes. Take-home naloxone is a key public health response to the overdose crisis where people who may witness an overdose are educated on naloxone administration and overdose response. This study aims to investigate signs and substances associated with naloxone administration stratified by opioid contribution to death and substances detected where naloxone was administered but opioids did not contribute to death.

This observational study of Canadian coroner and medical examiner files from 2016 to 2017 uses descriptive and chi square analyses to determine what signs were associated with having been administered naloxone among people for whom opioids contributed to death and what substances were associated with naloxone administration among people for whom no opioids were detected or contributed to the death.

The study included 1,521 people who died of acute drug toxicity. Naloxone was administered to 45% of those who died and was administered in higher proportions when opioids contributed to death (50%) compared to when opioids did not (35%). Among people for whom opioids contributed to death, having been administered naloxone was associated with expected opioid toxicity signs of snoring/gurgling, altered consciousness, appearing asleep, and being cold. Among people for whom opioids did not contribute to death, detection of opioids or stimulants were significantly associated with naloxone administration.

Further investigation into the barriers to naloxone administration should be conducted in order to improve drug toxicity response education.

## Introduction

North America has witnessed a crisis of high numbers of drug toxicity deaths (also referred to as overdoses or drug poisoning deaths) in the last decade, mainly attributable to opioids [1, 2]. Opioids impart an analgesic and euphoric effect which predominantly manifest when the drug class acts on µ receptors in the brain and spinal cord [3]. Opioid toxicity occurs when someone takes a higher dose of opioids than their body can handle, which causes respiratory depression leading to hypoxemia and hypercapnia, and subsequently cardiac arrest, brain injury, and death [3].

Naloxone is a reverse opioid agonist which binds to µ receptors, displaces opioids, and can temporarily reverse signs and symptoms associated with opioid toxicity [3]. The effects of naloxone are specific to opioids; naloxone is not a treatment for toxicity caused by stimulants, benzodiazepines, or alcohol [3]. Take-home naloxone programs distribute naloxone to individuals to use if they witness an opioid toxicity event and provide education on how to respond in a drug toxicity event [4]. Take-home naloxone programs are quantitatively demonstrated to save lives [5, 6]. Between April 2016 and December 2017 an estimated 1,580 death events were prevented by take-home naloxone programs in British Columbia alone [5]. Overdose education and naloxone distribution programs provide the education needed to effectively respond to an overdose [6].

Publicly funded take-home naloxone programs operate in all provinces and territories in Canada [4]. Provincial take-home naloxone programs existed in British Columbia, Ontario, Saskatchewan and Alberta prior to 2016. During 2016, Newfoundland and Labrador and the Northwest Territories launched take-home naloxone programs. And during 2017, Manitoba, the Yukon, Nunavut, Nova Scotia, Prince Edward Island, New Brunswick and Quebec launched take-home naloxone programs [4]. However, inconsistent guidance on how to identify and respond to drug toxicity currently presents challenges [7]. One important aspect of overdose response education is to use a common set of signs and symptoms to determine response [7].

A toxidrome is a grouping of commonly presenting physical findings that are used to assess and treat drug toxicity [8]. The opioid toxidrome includes decreased respiratory rate and blood pressure, constricted pupils and depressed mental status [8]. The Government of Canada has provided guidance on opioid toxicity, highlighting important signs of an opioid overdose. These include trouble walking or talking, blue or gray lips or nails, very small pupils, cold and damp skin, dizziness, confusion, extreme sleepiness, choking or gurgling sounds, slow or weak breathing, and being unable to wake up even when shaken or shouted at [9].

Responding to drug toxicity becomes more challenging when multiple substances are involved and produce a mix of signs and symptoms from different toxidromes [10]. Currently in Canada, multi-drug toxicity contributes to most drug toxicity deaths; 65% of accidental apparent opioid toxicity deaths occurring from January to June of 2024 also involved a stimulant, and the same proportion involved some other psychoactive substance [2]. The illegal drug supply in Canada is dominated by mixed products containing multiple classes of drugs in inconsistent ratios, which presents additional challenges when responding to drug toxicity as people may not know what they have consumed [11]. For example, the co-consumption of opioids with substances such as benzodiazepines and benzodiazepine analogues complicates the resuscitation of people experiencing opioid toxicity as naloxone can only reverse the effects of opioids [10]. Drug checking services have identified potent fentanyl analogues, benzodiazepines and non-opioid tranquilizers like medetomidine and xylazine in the Canadian opioid supply [12, 13].

Using coroner and medical examiner data, we aimed to understand the associations between toxicity signs observed, naloxone administration, and the substances detected on toxicology reports for people who died of drug toxicity. First, we describe the signs observed when naloxone was administered to people for whom opioids contributed or did not contribute to death. Second, we examine what substance classes and substance class combinations were detected on toxicology when naloxone was administered, but no opioids were detected or contributed to the death.

## Methods

### Data Source

We used data from a chart review of coroner and medical examiner records of substance-related acute toxicity deaths in Canada for individuals who died between January 1, 2016 and December 31, 2017 [14]. Individuals were considered to have died due to acute toxicity if death was determined to be caused by the direct effects of the administration of exogenous substances, where at least one substance was either a drug or alcohol [14]. The study protocol and general description of the dataset are published elsewhere [14, 15].

#### Study Population

This study included people who died from acute drug toxicity in 2016 or 2017 in Canada whose death was witnessed, who had observed signs and symptoms recorded, and who had information on whether naloxone was administered. People who were missing information for study variables were excluded from the study population (Fig 1).

**Figure 1.**
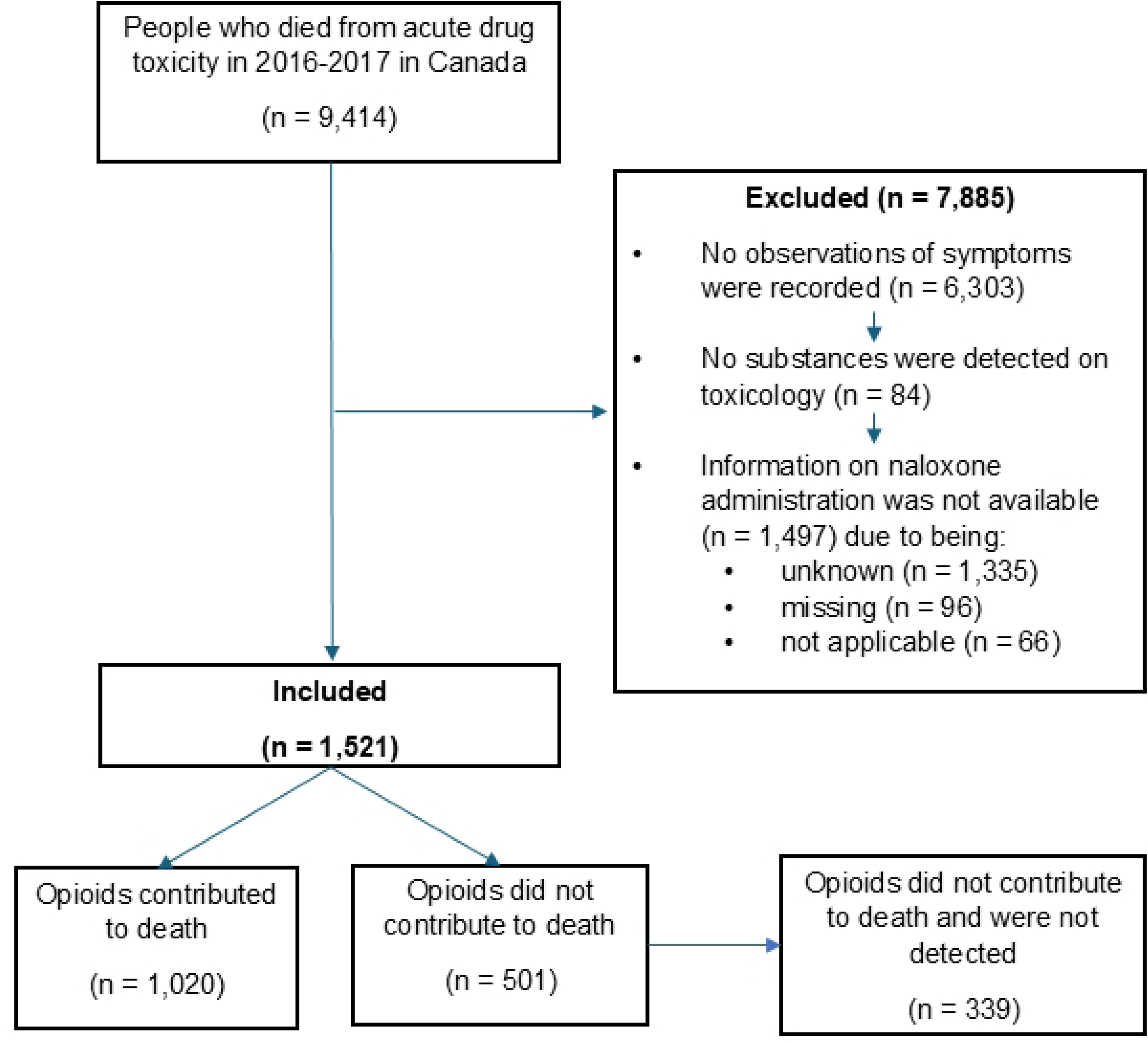
Flowchart for study population selection. Note: Number of included and excluded does not equal the entire sample due to random rounding. Categories for exclusion from the study sample are not mutually exclusive. Categories for exclusion were reviewed sequentially, therefore these are not total counts of people missing each criterion.

#### Variables

Key variables for these analyses included naloxone administration (administered by any person: bystander, first responder, emergency medical services, or hospital staff), witnessed signs (noted in the coroner and medical examiners and witnessed by any person such as bystanders, first responders, emergency medical services, or hospital staff), substance class contributing to death (where the direct effects of the substance led to death as determined by the coroner or medical examiner), and substance class detected (where the substance was present in toxicology testing and may or may not have contributed to death). Data were collected from coroner and medical examiner files by trained data abstractors using a standardized data collection tool. The standardized data collection tool included an open text option for other observed signs. CH reviewed open text responses, reclassified answers into existing sign categories, and generated new sign categories where necessary. Reclassification decisions were reviewed by authors AV and MF and consensus was reached within the research team. See Supplementary Table 1 for a grouping of substances by class. Opioids were grouped as a single class.

### Analyses

We investigated the reported numbers and proportion of people who reportedly exhibited signs and symptoms stratified by whether an opioid did or did not contribute to death and whether naloxone was administered. The substances detected among people for whom opioids neither contributed to death nor were detected and who received naloxone were quantified. Chi square tests were calculated based on naloxone administration. An UpSet plot was created to illustrate the combinations of substance classes detected among people who were administered naloxone despite an opioid not contributing to their death. To protect anonymity, the plot only shows substance class combinations detected in ten or more people.

As the study period coincided with many Canadian territories and provinces launching take-home naloxone programs, analyses were repeated with only deaths occurring in provinces with pre-existing programs (British Columbia, Ontario, Saskatchewan and Alberta). We conducted this analysis to determine whether associations were the same in different conditions.

Statistical analyses were completed in RStudio (Version 2022.02.0+443) and the UpSet plot was created using the ComplexUpset package [16, 17]. The STROBE statement cross-sectional checklist was used to guide project reporting [18].

#### Privacy and Ethics

All counts were randomly rounded to base three and percentages are calculated from these rounded numbers [19]. Counts less than 10, and percentages based on these counts, are suppressed. As chi square test statistics and exact p-values are based on the original counts, they are not shared to protect random rounding.

This study was reviewed and approved by the Health Canada and Public Health Agency of Canada Research Ethics Board (REB 2018-027P), the University of Manitoba Health Research Ethics Board (HS22710), and Newfoundland and Labrador Health Research Ethics Board (20200153).

## Results

A total of 1,521 people who died met study inclusion criteria. Opioids did not contribute to the deaths of approximately a third of the people included in the sample (n = 501). Figure 1 presents a flowchart of inclusion and exclusion in this analysis.

### Naloxone Administration

Where naloxone administration was known, naloxone was administered in just under half of all deaths with witnessed signs (n = 687, 45% of those with known naloxone administration). Half of people for whom opioids contributed to death were administered naloxone (n = 513, 50%), while 35% of people for whom opioids did not contribute to death received naloxone (n = 174).

Among people for whom an opioid contributed to death, naloxone administration was significantly associated with observed signs of altered consciousness, appearing asleep, feeling cold, and snoring/gurgling (Table 1).

**Table 1.**
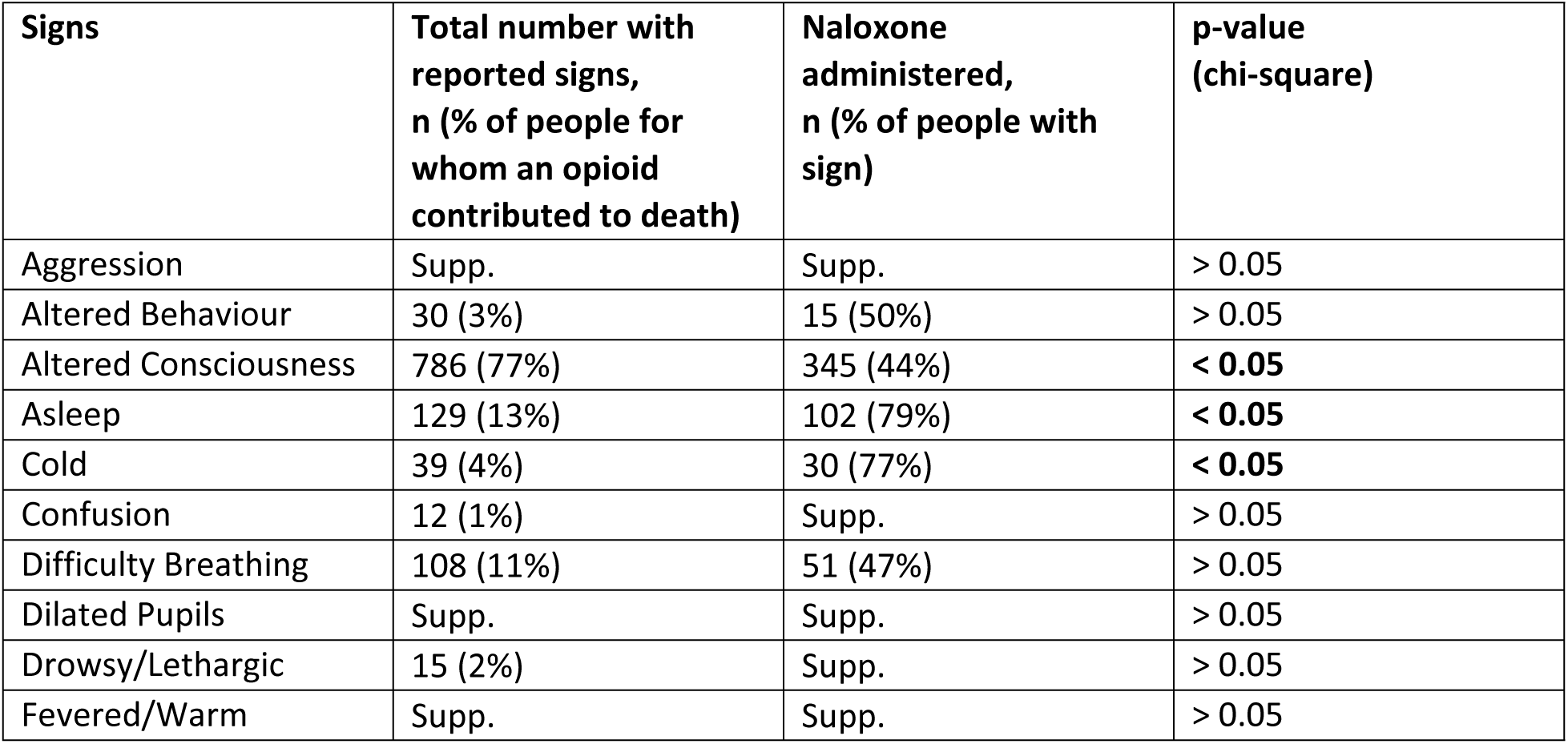
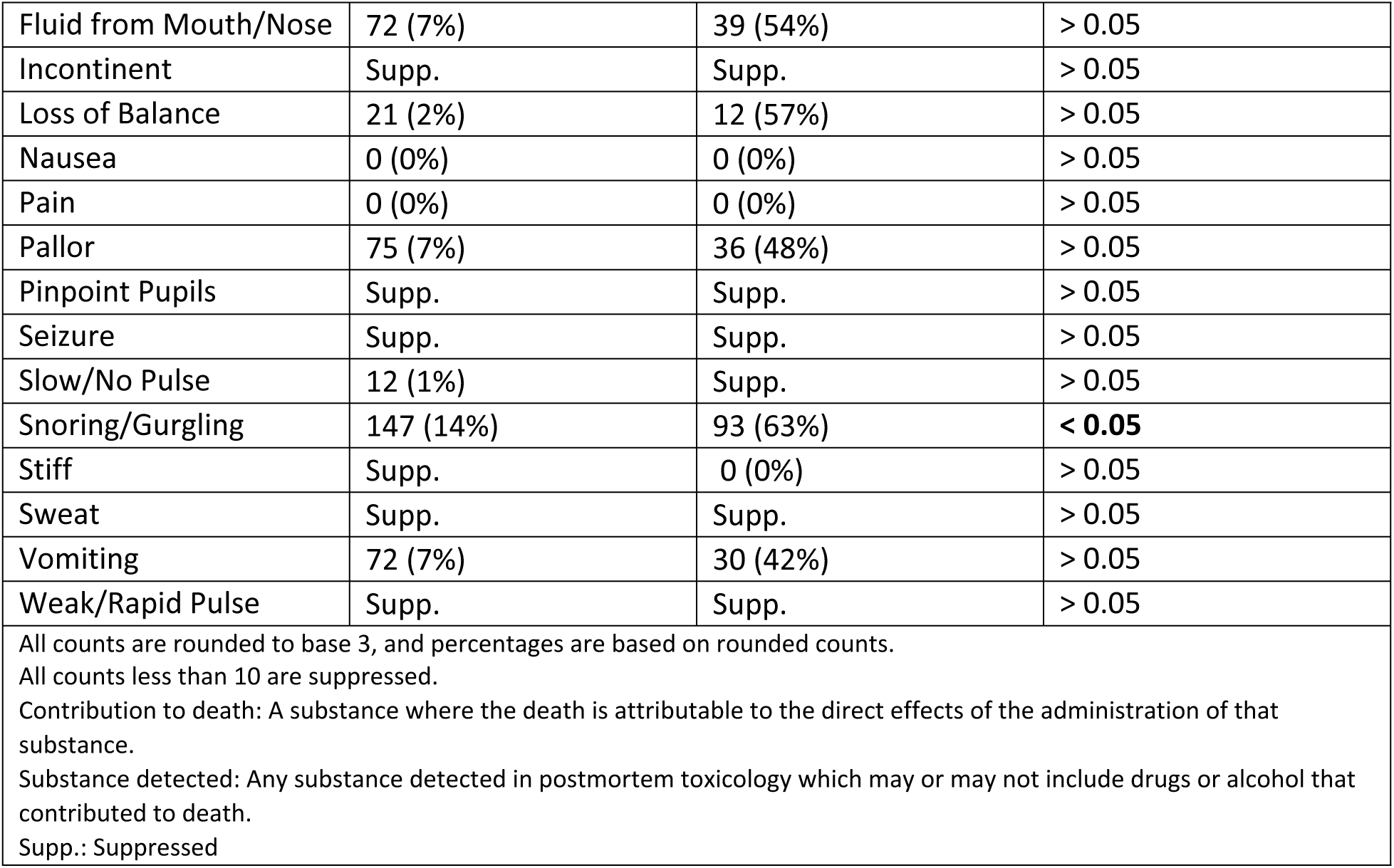
Naloxone administration by observed signs among people who died of acute toxicity for whom an opioid contributed to death, Canada, 2016-2017 (N = 1020).

| Signs | Total number with reported signs, n (% of people for whom an opioid contributed to death) | Naloxone administered, n (% of people with sign) | p-value (chi-square) |
| --- | --- | --- | --- |
| Aggression | Supp. | Supp. | > 0.05 |
| Altered Behaviour | 30 (3%) | 15 (50%) | > 0.05 |
| Altered Consciousness | 786 (77%) | 345 (44%) | < **0.05** |
| Asleep | 129 (13%) | 102 (79%) | < **0.05** |
| Cold | 39 (4%) | 30 (77%) | < **0.05** |
| Confusion | 12 (1%) | Supp. | > 0.05 |
| Difficulty Breathing | 108 (11%) | 51 (47%) | > 0.05 |
| Dilated Pupils | Supp. | Supp. | > 0.05 |
| Drowsy/Lethargic | 15 (2%) | Supp. | > 0.05 |
| Fevered/Warm | Supp. | Supp. | > 0.05 |
| Fluid from Mouth/Nose | 72 (7%) | 39 (54%) | > 0.05 |
| Incontinent | Supp. | Supp. | > 0.05 |
| Loss of Balance | 21 (2%) | 12 (57%) | > 0.05 |
| Nausea | 0 (0%) | 0 (0%) | > 0.05 |
| Pain | 0 (0%) | 0 (0%) | > 0.05 |
| Pallor | 75 (7%) | 36 (48%) | > 0.05 |
| Pinpoint Pupils | Supp. | Supp. | > 0.05 |
| Seizure | Supp. | Supp. | > 0.05 |
| Slow/No Pulse | 12 (1%) | Supp. | > 0.05 |
| Snoring/Gurgling | 147 (14%) | 93 (63%) | < **0.05** |
| Stiff | Supp. | 0 (0%) | > 0.05 |
| Sweat | Supp. | Supp. | > 0.05 |
| Vomiting | 72 (7%) | 30 (42%) | > 0.05 |
| Weak/Rapid Pulse | Supp. | Supp. | > 0.05 |
| All counts are rounded to base 3, and percentages are based on rounded counts.<br>All counts less than 10 are suppressed.<br>Contribution to death: A substance where the death is attributable to the direct effects of the administration of that substance.<br>Substance detected: Any substance detected in postmortem toxicology which may or may not include drugs or alcohol that contributed to death.<br>Supp.: Suppressed |  |  |  |

Among people for whom an opioid did not contribute to death, naloxone administration was significantly associated with detection of opioids or stimulants (Table 2).

**Table 2.**
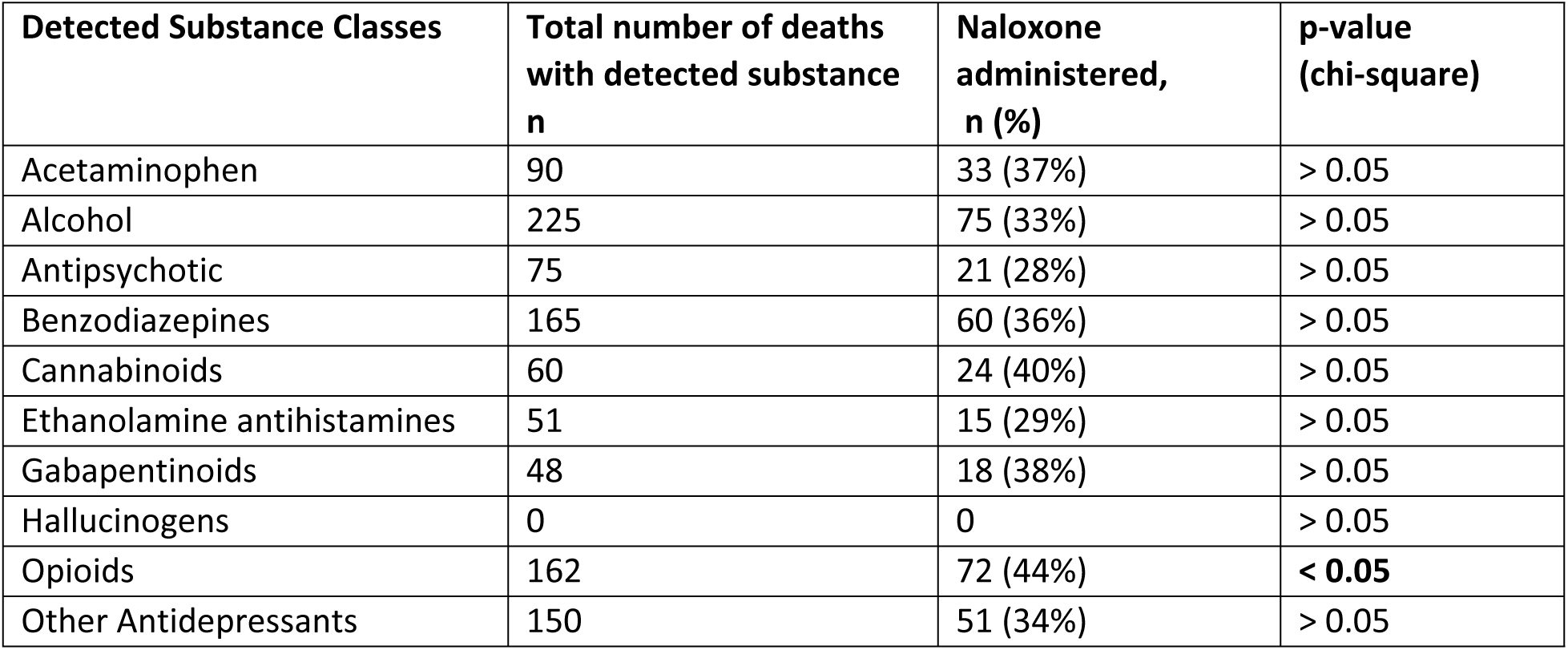
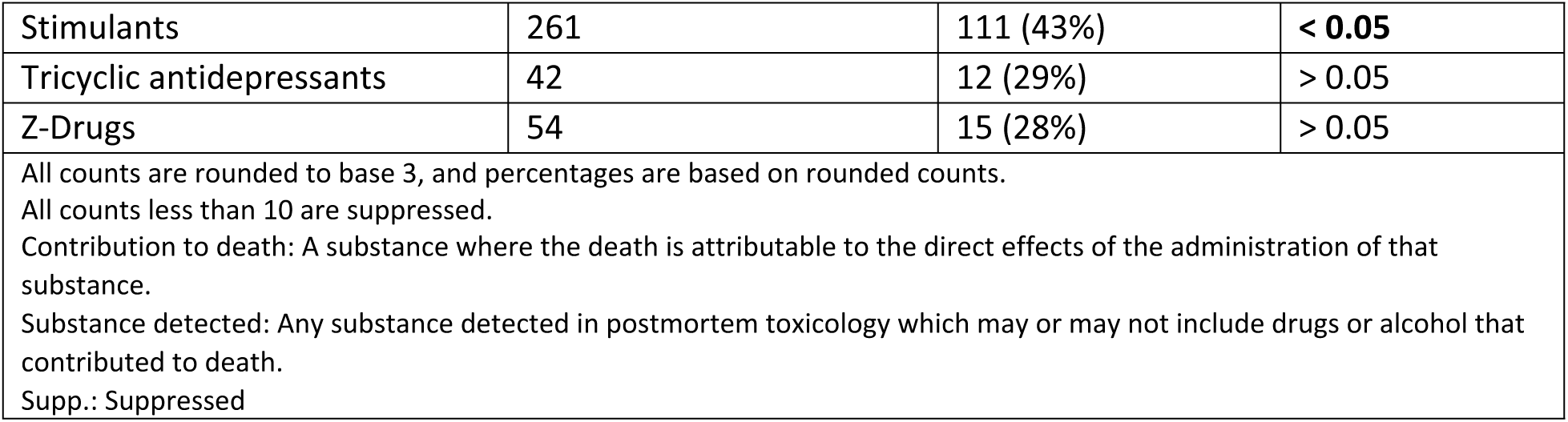
Naloxone administration by detected substance class among people who died of acute toxicity for whom opioids did not contribute to death, Canada, 2016-2017 (N = 501).

| Detected Substance Classes | Total number of deaths with detected substance n | Naloxone administered, n (%) | p-value (chi-square) |
| --- | --- | --- | --- |
| Acetaminophen | 90 | 33 (37%) | > 0.05 |
| Alcohol | 225 | 75 (33%) | > 0.05 |
| Antipsychotic | 75 | 21 (28%) | > 0.05 |
| Benzodiazepines | 165 | 60 (36%) | > 0.05 |
| Cannabinoids | 60 | 24 (40%) | > 0.05 |
| Ethanolamine antihistamines | 51 | 15 (29%) | > 0.05 |
| Gabapentinoids | 48 | 18 (38%) | > 0.05 |
| Hallucinogens | 0 | 0 | > 0.05 |
| Opioids | 162 | 72 (44%) | < **0.05** |
| Other Antidepressants | 150 | 51 (34%) | > 0.05 |
| Stimulants | 261 | 111 (43%) | < 0.05 |
| Tricyclic antidepressants | 42 | 12 (29%) | > 0.05 |
| Z-Drugs | 54 | 15 (28%) | > 0.05 |
| All counts are rounded to base 3, and percentages are based on rounded counts.<br>All counts less than 10 are suppressed.<br>Contribution to death: A substance where the death is attributable to the direct effects of the administration of that substance.<br>Substance detected: Any substance detected in postmortem toxicology which may or may not include drugs or alcohol that contributed to death.<br>Supp.: Suppressed |  |  |  |

Among people for whom opioids were not detected and did not contribute to death, only the presence of stimulants was significantly associated with naloxone administration (Table 3).

**Table 3.**
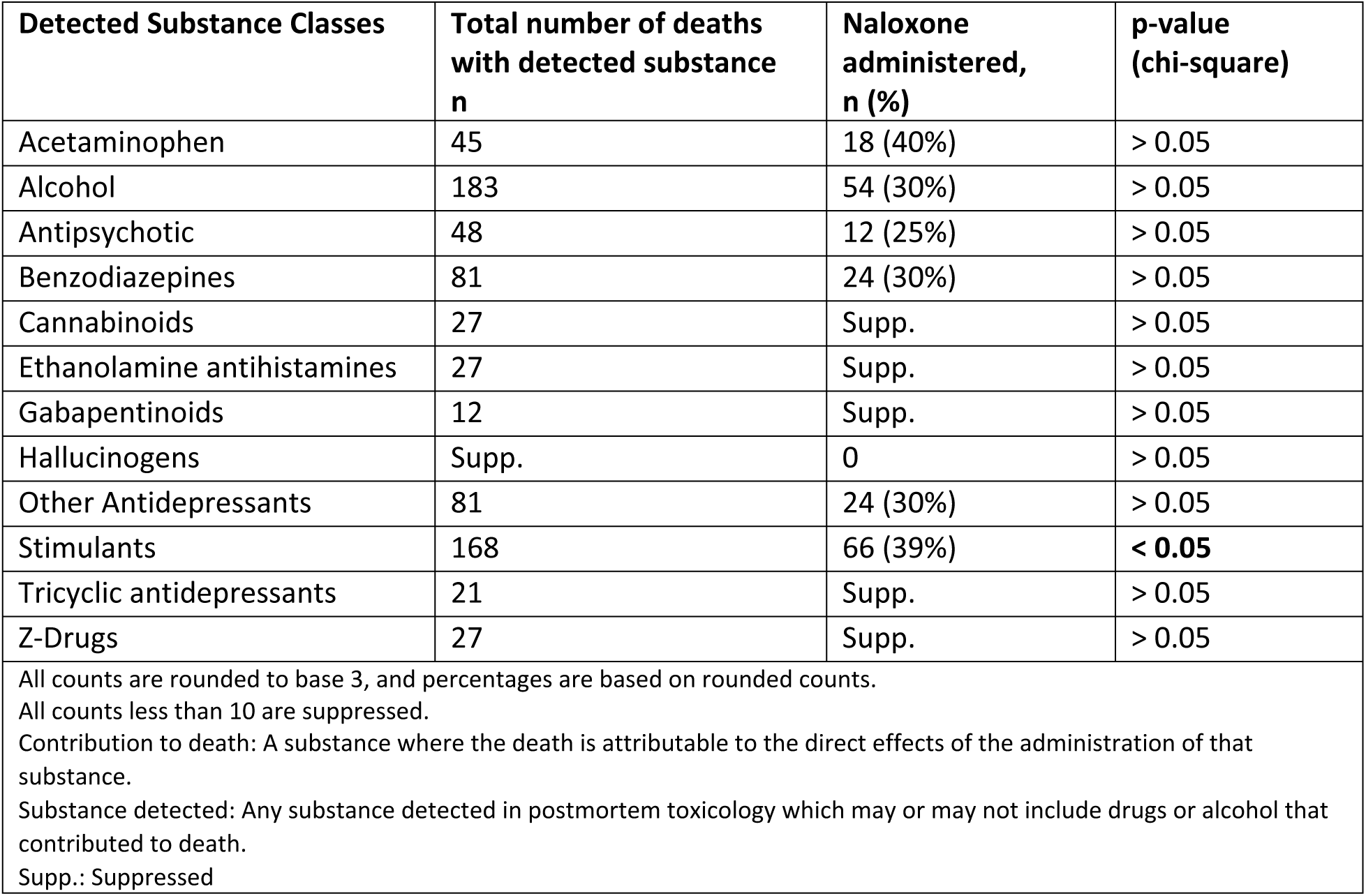
Naloxone administration by detected substance class among people for whom opioids did not contribute to the death and were not detected in toxicology, Canada, 2016-2017 (N = 339).

| Detected Substance Classes | Total number of deaths with detected substance<br>n | Naloxone administered,<br>n (%) | p-value<br>(chi-square) |
| --- | --- | --- | --- |
| Acetaminophen | 45 | 18 (40%) | > 0.05 |
| Alcohol | 183 | 54 (30%) | > 0.05 |
| Antipsychotic | 48 | 12 (25%) | > 0.05 |
| Benzodiazepines | 81 | 24 (30%) | > 0.05 |
| Cannabinoids | 27 | Supp. | > 0.05 |
| Ethanolamine antihistamines | 27 | Supp. | > 0.05 |
| Gabapentinoids | 12 | Supp. | > 0.05 |
| Hallucinogens | Supp. | 0 | > 0.05 |
| Other Antidepressants | 81 | 24 (30%) | > 0.05 |
| Stimulants | 168 | 66 (39%) | < 0.05 |
| Tricyclic antidepressants | 21 | Supp. | > 0.05 |
| Z-Drugs | 27 | Supp. | > 0.05 |
| All counts are rounded to base 3, and percentages are based on rounded counts.<br>All counts less than 10 are suppressed.<br>Contribution to death: A substance where the death is attributable to the direct effects of the administration of that substance.<br>Substance detected: Any substance detected in postmortem toxicology which may or may not include drugs or alcohol that contributed to death.<br>Supp.: Suppressed |  |  |  |

The most frequently observed combinations of drug classes among people for whom opioids did not contribute to death and were administered naloxone include stimulants and alcohol (n = 45), stimulants and opioids (n = 45), and opioids and benzodiazepines (n = 33) (Fig 2).

**Figure 2.**
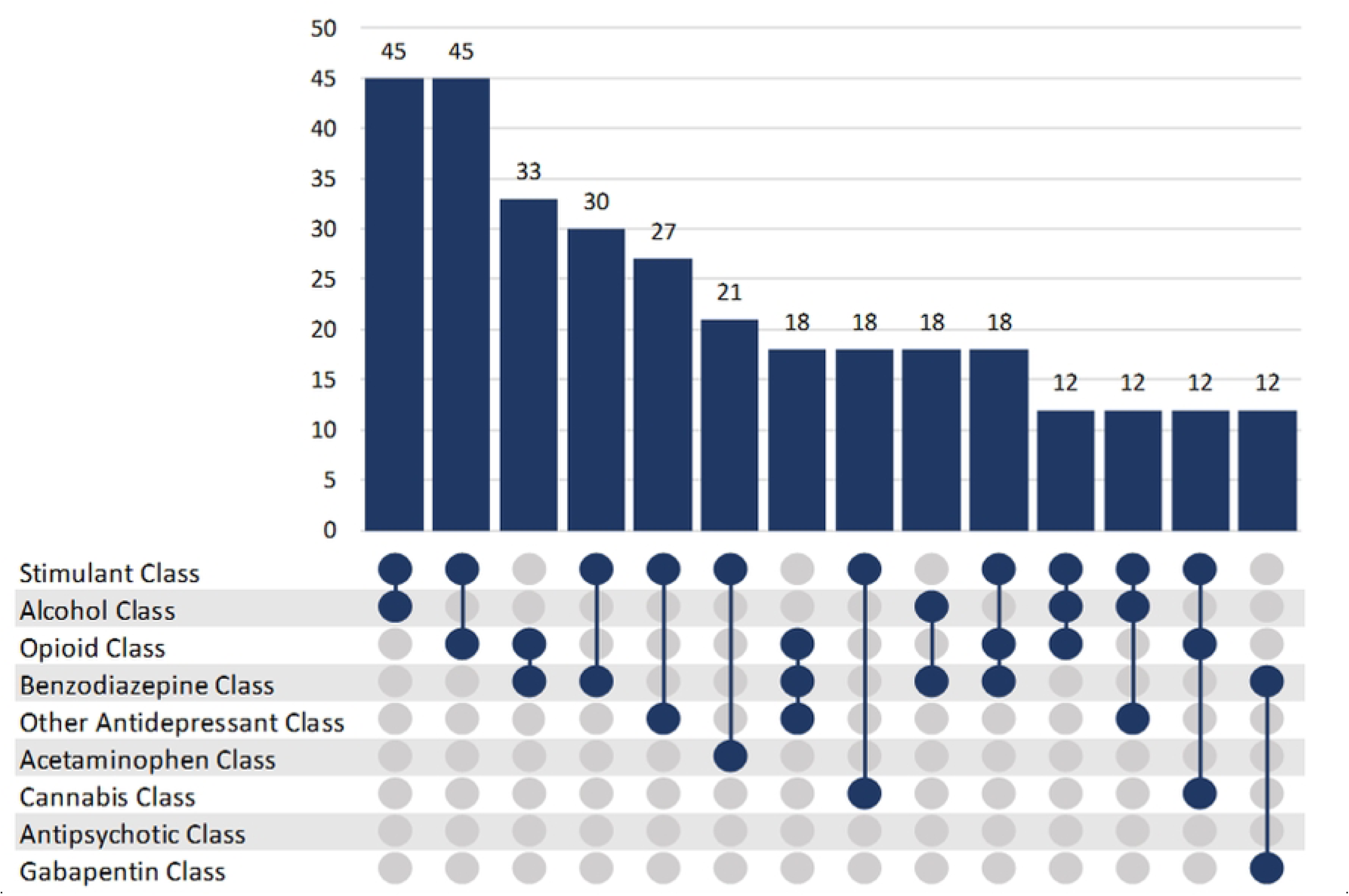
Substance class combinations detected in more than ten individuals among people who died of acute toxicity for whom opioids did not contribute to death and naloxone was administered, Canada, 2016-2017. Dots indicate a substance class that was detected, with lines connecting co-detected substance classes. Frequencies of these substance class combinations are indicated by the bars. As these combinations are inclusive, other substances may have also been detected.

Supplementary Tables 2, 3 and 4 show the same analyses conducted with only the provinces with publicly funded take-home naloxone programs established prior to the study period (British Columbia, Ontario, Saskatchewan and Alberta) to determine whether results would be different in jurisdictions with greater availability of naloxone. There were no differences in significance of associations in the analysis limited by jurisdictions with publicly funded take-home naloxone programs compared to the entire sample in all Canadian jurisdictions.

## Discussion

Where witness observations were available, naloxone was administered in only half of people for whom opioids contributed to death. Low proportions of naloxone administration appear in other jurisdictions. An analysis of 2019 data from 37 states and the District of Columbia in the United States of America found no evidence of naloxone administration in 77.3% of opioid involved overdose deaths [20]. The findings from Quinn et al. paper and our analysis are only based on people who have died offering a potential explanation for the low proportion of naloxone detected and underscoring the importance of the intervention. As naloxone is such an important and lifesaving intervention, our finding prompts investigation into the impediments to naloxone administration in Canada, whether low proportion of naloxone administration persists, and strategies that might improve these figures.

Changes in legal protection that occurred during the study period can inform the finding of low rates of naloxone administration. In Canada, the *Good Samaritan Drug Overdose Act* was enacted in May 2017 to protect those responding to overdoses against charges for possession of a controlled substance or breaches of conditions regarding simple possession of controlled substances [21]. While the *Good Samaritan Drug Overdose Act* would have only been in effect for a portion of the study period, it may have impacted the willingness of bystanders to engage in an overdose response in data collected from May to December 2017. However, the impact of the Act may have been limited as knowledge of the *Act* remained low [22].

The signs associated with naloxone administration in people for whom opioids contributed to death included altered consciousness, appearing asleep, feeling cold, and snoring/gurgling. Witness reports of altered consciousness or appearing asleep is likely consistent with the toxidromal concept of “depressed mental status”, although it can be difficult to differentiate between sleep and altered level of consciousness for witnesses with limited experience with drug toxicity [8]. Snoring or gurgling is associated with respiratory depression and is caused by opioid, benzodiazepine, alcohol toxicity either independently or additively [9]. Feeling cold may have occurred before or after death and therefore some of the people who were observed to feel cold may have already died when naloxone was administered. Notably, while constricted pupils are a commonly cited sign of opioid toxicity, fewer than 10 people had pinpoint pupils noted in their charts [8, 9]. It is possible that some witnesses did not know to check for this sign, and it is also possible that this sign is of limited clinical utility. Our study suggests that conventional toxidromes are of limited utility in assessing the underlying cause of toxicological emergencies that may proceed to death; while many signs of opioid toxicity were evident among people for whom an opioid contributed to death, they were not reliably present or consistently noticed by witnesses.

The number of people exhibiting aggression was suppressed due to insufficient case counts. The assumption that people who use drugs will exhibit aggression after receiving naloxone plays into the stereotype of people who use drugs as violent and antisocial [23] and may effect bystander willingness to administer naloxone [24]. While the people in this study died from the toxicity event they experienced it is notable that altered behaviour was reported in sufficiently high numbers to be reported whereas aggression was not. A study conducted by Neale et al. found that people who experienced opioid toxicity were more likely to display anger if the person resuscitating them criticized, berated, or chastised them and were less likely to display anger if the person resuscitating them communicated positively with them while withdrawal symptoms were not associated with displays of anger [24].

Naloxone administration is also associated with stimulant detection. While signs associated with stimulant toxicity are typically distinct from those associated with opioid toxicity, we see in Figure 2 that the most common combinations with stimulants include alcohol, opioids, and benzodiazepines, which are all depressants and may share some signs and symptoms with the opioid toxidrome. Signs such as confusion, drowsiness, slurred speech, and loss of coordination and balance are apparent in use of alcohol, benzodiazepines, as well as opioids [9, 25, 26]. The signs that led the responder to administer naloxone may have been caused by a co-detected substance. There might also be socio-demographic differences and different social networks between people who use stimulants and people who use other drugs aside from stimulants or opioids that may result in more community naloxone knowledge, availability, and administration. The social contexts of using different drugs and their effect on naloxone administration could be an avenue for future investigation.

People for whom opioids did not contribute to death were not more likely to have been administered naloxone when a benzodiazepine was detected. When benzodiazepines are mixed with opioids, benzodiazepine-related sedation may cause confusion in overdose response and incomplete response to naloxone [10]. While sedation related to benzodiazepines might be expected to be associated with naloxone administration, this was not apparent in our analyses. It may be that concentrations of benzodiazepines were not sufficiently high to cause signs and symptoms during the study period. Benzodiazepines became more prevalent in the illegal drug supply in the years that followed [10, 27, 28]. Health Canada’s Drug Analysis Service detected benzodiazepines more frequently in illegal substances seized by Canadian law enforcement agencies, particularly in samples that also contained opioids [27, 28]. Of the people who died from opioid toxicity in 2024, 34% also had a benzodiazepine contribute to their death, compared to 8% in 2018 [28]. It’s unclear whether the climbing prevalence of benzodiazepines may result in increased administration of naloxone in the absence of opioid toxicity.

Concurrent use of different substances can complicate how people responding to drug toxicity determine whether to administer naloxone due to unusual symptomology. People may use different substances to counteract or balance the subjective experience of a drug, enhance that experience, mimic the effect of an unavailable or expensive substance, or to self-medicate for a pre-existing condition; many people may also unknowingly use substances which are adulterating the drugs they aim to use [10, 29, 30]. As concurrent use of substances is likely to continue, take-home naloxone programs in Canada will benefit from a shared understanding on drug toxicity that reflects the changing nature of the illegal drug supply; and pan-Canadian guidance on response [7]. Canadian harm reduction and take-home naloxone programs are working to adapt education around drug toxicity response [31]. For example, in contrast to the conventional opioid toxidrome, as of 2023 guidance from British Columbia directs overdose responders to evaluate respiratory status as the primary factor in whether to administer naloxone versus a cluster of signs [31]. There is an appetite for increased education on naloxone administration across the country; a Canadian guidance development project identified the desire for standardized education on naloxone administration based on community feedback [7].

### Limitations

The data included in this analysis were extracted from coroner and medical examiner charts; the purpose of these charts is to report on a death investigation and may underreport or miss data on bystander and first responder observed signs. Appearing asleep and having altered level of consciousness might not have been distinguishable for many bystanders. The data set includes only fatal acute toxicity events and perhaps only more severe or late-stage toxicity signs may have been observed as a result. The signs and symptoms observed during fatal events may differ from non-fatal events, however, data on signs and symptoms of toxicity observed in the community are difficult to acquire.

The chart review study collected data on whether a substance contributed to death or was detected in toxicology reports; but did not collect information on the quantities of the substances detected in toxicology reports. It is not possible to determine whether detected substances were present in sufficient quantities to cause signs of toxicity (and these quantities could vary based on individual factors).

## Conclusion

The proportion of naloxone administration among people for whom opioids contributed to death was low. During fatal acute toxicity events where an opioid contributed to death, naloxone was most often administered to people who were observed to be asleep, have altered consciousness, be cold to the touch, and/or be snoring or gurgling. When naloxone was administered to people who did not have an opioid contribute to their death, depressive substances were often detected and potentially mimicking opioid toxicity signs. Further investigation into factors inhibiting naloxone administration is needed to improve programs and save lives.

## Data Availability

The national chart review of coroner and medical examiner files dataset generated and analyzed for the current study is not publicly available due to provisions in the data use agreements with provincial and territorial data providers. The corresponding author can assist in directing inquiries about data access to the original data providers.

## Acknowledgements

We would like to acknowledge our collaborators at the offices of chief coroners and chief medical examiners across Canada for providing access to their death investigation files, and Jenny Rotondo, Brandi Abele, Songul Bozat-Emre, Matthew Bowes, Jessica Halverson, Dirk Huyer, Beth Jackson, Graham Jones, Jennifer Leason, Regan Murray, Erin Rees and Emily Schleihauf for their contributions to developing the National Chart Review Study of Substance-Related Acute Toxicity Deaths. Thanks to Aganeta Enns for help with formatting.

## Supporting information

**Supplementary Table 1. List of substances by substance type.**

**Supplementary Table 2. Repeated analyses from Table 1 in provinces with existing take-home naloxone programs (British Columbia, Ontario, Saskatchewan and Alberta): Naloxone administration by observed signs among people who died of acute toxicity where an opioid contributed to the death, Canada, 2016-2017 (N = 903)**

**Supplementary Table 3. Repeated analyses from Table 2 in provinces with existing take-home naloxone programs (British Columbia, Ontario, Saskatchewan and Alberta): Naloxone administration by detected substance class among people who died of acute toxicity where opioids did not contribute to the death, Canada, 2016-2017 (N = 903)**

**Supplementary Table 4. Repeated analyses from Table 3 in provinces with existing take-home naloxone programs (British Columbia, Ontario, Saskatchewan and Alberta): Naloxone administration by detected substance class among people who died of acute toxicity where opioids did not contribute to the death and were not detected in toxicology, Canada, 2016-2017 (N = 255)**

## References

1. Ahmad FB, Cisewski JA, Rossen LM, Sutton P. Provisional Drug Overdose Death Counts. National Center for Health Statistics; 2026. Available from: https://www.cdc.gov/nchs/nvss/vsrr/drug-overdose-data.htm

2. Federal provincial and territorial Special Advisory Committee on the Epidemic of Opioid Overdoses. Opioid- and Stimulant-related Harms in Canada. Ottawa: Public Health Agency of Canada; June 2026. Available from: https://health-infobase.canada.ca/substance-related-harms/opioids-stimulants/

3. Dezfulian C, Orkin AM, Maron BA, Elmer J, Girotra S, Gladwin MT, et al. Opioid-associated out-of-hospital cardiac arrest: Distinctive clinical features and implications for health care and public responses: A scientific statement from the American Heart Association. Circulation. 2021;143(16):e836–e70. 10.1161/CIR.0000000000000958

4. Moustaqim-Barrette A, Elton-Marshall T, Leece P, Morissette C, Rittenbach K, Buxton J. Environmental scan: Naloxone access and distribution in Canada. Vancouver: Canadian Research Initiative in Substance Misuse (CRISM); 2019. Available from: https://crism.ca/2019/06/13/naloxone-distribution-environmental-scan/

5. Irvine MA, Kuo M, Buxton JA, Balshaw R, Otterstatter M, Macdougall L, et al. Modelling the combined impact of interventions in averting deaths during a synthetic-opioid overdose epidemic. Addiction. 2019;114(9):1602–13. 10.1111/add.14664

6. Razaghizad A, Windle SB, Filion KB, Gore G, Kudrina I, Paraskevopoulos E, et al. The effect of overdose education and naloxone distribution: An umbrella review of systematic reviews. Am J Public Health. 2021;111(8):e1–e12. 10.2105/AJPH.2021.306306

7. Ferguson M, Rittenbach K, Leece P, Adams A, Ali F, Elton-Marshall T, et al. Guidance on take-home naloxone distribution and use by community overdose responders in Canada. CMAJ. 2023;195(33):E1112–E23. 10.1503/cmaj.230128

8. Brocato C, Paley RJ. Toxidromes: Common poisoning syndromes to know. Journal of Emergency Medical Services. 2021. Available from: https://www.jems.com/patient-care/emergency-medical-care/common-poisoning-syndromes-to-know/

9. Opioid overdose. Ottawa: Health Canada; 2023. Available from: https://www.canada.ca/en/health-canada/services/opioids/overdose.html

10. Purssell R, Buxton J, Godwin J, Moe J. Potent sedatives in opioids in BC: Implications for resuscitation, and benzodiazepine and etizolam withdrawal. BC Medical Journal 2021;63(4):177–8. Available from: https://bcmj.org/bccdc/potent-sedatives-opioids-bc-implications-resuscitation-and-benzodiazepine-and-etizolam

11. Canadian Centre on Substance Use and Addiction. Community urinalysis and self-report project: Cross-Canada report on the use of drugs from the unregulated supply, 2019-2021 data. Ottawa, Ont.: CCSA; 2022. Available from: https://www.ccsa.ca/en/community-urinalysis-and-self-report-project-cross-canada-report-use-drugs-unregulated-supply-2019

12. Toronto Drug Checking Service. What’s in Toronto’s drug supply? 2024. Available from: https://drugchecking.community/.

13. British Columbia Centre on Substance Use. Drug checking results 2024. Available from: https://bccsu-drugsense.onrender.com/.

14. Public Health Agency of Canada. Substance-related acute toxicity deaths in Canada from 2016 to 2017: A review of coroner and medical examiner findings. Ottawa: PHAC; 2022. Available from: https://www.canada.ca/en/health-canada/services/opioids/data-surveillance-research/substance-related-acute-toxicity-deaths-canada-2016-2017-review-coroner-medical-examiner-files.html

15. Rotondo J, VanSteelandt A, Kouyoumdjian F, Bowes MJ, Kakkar T, Jones G, et al. Substance-Related Acute Toxicity Deaths in Canada from 2016 to 2017: Protocol for a Retrospective Chart Review Study of Coroner and Medical Examiner Files. JMIR Public Health and Surveillance. 2025;11:e49981. 10.2196/49981

16. Krassowski M. ComplexUpset. Zenodo; 2020. 10.5281/zenodo.3700590

17. R Core Team. R: A Language and Environment for Statistical Computing. Vienna, Austria: R Foundation for Statistical Computing; 2022. https://www.r-project.org/

18. Cuschieri S. The STROBE guidelines. Saudi J Anaesth. 2019;13(Suppl 1):S31–S4. 10.4103/sja.sja_543_18

19. VanSteelandt A, Chang GY, McKenzie K, Kouyoumdjian F. Accidental substance-related acute toxicity deaths among youth in Canada: a descriptive analysis of a national chart review study of coroner and medical examiner data. Health Promot Chronic Dis Prev Can. 2024;44(3):77–88. 10.24095/hpcdp.44.3.02

20. Quinn K, Kumar S, Hunter CT, O’Donnell J, Davis NL. Naloxone administration among opioid-involved overdose deaths in 38 United States jurisdictions in the State Unintentional Drug Overdose Reporting System, 2019. Drug Alcohol Depend. 2022;235:109467. 10.1016/j.drugalcdep.2022.109467

21. Government of Canada. About the Good Samaritan Drug Overdose Act. Ottawa: Health Canada; 2021. Available from: https://www.canada.ca/en/health-canada/services/opioids/about-good-samaritan-drug-overdose-act.html.

22. Kievit B, Xavier JC, Ferguson M, Palis H, Moallef S, Slaunwhite A, et al. Intention to seek emergency medical services during community overdose events in British Columbia, Canada: a cross-sectional survey. Subst Abuse Treat Prev Policy. 2022;17(1):56. 10.1186/s13011-022-00484-0

23. Ferguson N, Farrugia A, Moore D, Fraser S. Remaking the ‘angry Narcanned subject’: Affording new subject positions through take-home naloxone training. International Journal of Drug Policy. 2024;123. 10.1016/j.drugpo.2023.104253

24. Neale J, Kalk NJ, Parkin S, Brown C, Brandt L, Campbell ANC, et al. Factors associated with withdrawal symptoms and anger among people resuscitated from an opioid overdose by take-home naloxone: Exploratory mixed methods analysis. J Subst Abuse Treat. 2020;117:108099. 10.1016/j.jsat.2020.108099

25. Government of Canada. Health risks of alcohol. Ottawa: Health Canada; 2021. Available from: https://www.canada.ca/en/health-canada/services/substance-use/alcohol/health-risks.html.

26. Government of Canada. Benzodiazepines: Ottawa: Health Canada; 2023. Available from: https://www.canada.ca/en/health-canada/services/substance-use/controlled-illegal-drugs/benzodiazepines.html.

27. Government of Canada. Analyzed Drug Report. Longueuil (QC): Health Canada’s Drug Analysis Service (DAS) and Cannabis Laboratory (CL); 2024. Available from: https://health-infobase.canada.ca/drug-analysis-service/analyzed-drug-report.html

28. Federal provincial and territorial Special Advisory Committee on the Epidemic of Opioid Overdoses. Benzodiazepines in apparent drug toxicity deaths in Canada, 2018 to 2024. Ottawa: Public Health Agency of Canada; June 2026. Available from: https://health-infobase.canada.ca/substances/harms/benzodiazepines/

29. Boileau-Falardeau M, Contreras G, Gariepy G, Laprise C. Patterns and motivations of polysubstance use: a rapid review of the qualitative evidence. Health Promot Chronic Dis Prev Can. 2022;42(2):47–59. 10.24095/hpcdp.42.2.01

30. Hays HL, Spiller HA, DeRienz RT, Rine NI, Guo HT, Seidenfeld M, et al. Evaluation of the relationship of xylazine and fentanyl blood concentrations among fentanyl-associated fatalities. Clinical toxicology. 2024;62(1), 26–31. 10.1080/15563650.2024.2309326

31. BCCDC Harm Reduction Services. Responding to Prolonged Sedation. Towards the Heart; 2023. https://towardtheheart.com/assets/uploads/1740421710kl2TGBqtjoWPhYm4l86LWKwPXF3vRMm2sOTExNI.pdf

